## Supplementary Material for "Design Considerations for Technology-assisted Fall-Resisting Skills Training Trials in Older Adults: A Pilot and Feasibility Study"

**Table S1**. Overview of missing data per task.

| Task category | Task | Correct data (n) | Missing data (n) | Comments |
|---|---|---|---|---|
| Familiarisation | Walking at different speeds | 11 | 0 | |
| Non-task-specific balance tasks | Boat | 11 | 0 | |
| | Car | 11 | 0 | |
| | Walking with AST | 11 | 0 | |
| PGA | Obstacle avoidance | 8 | 3 | For 2 participants, only data of the first half due to technical issues |
| | Stepping stones – obstacles | 11 | 0 | |
| | Stepping stones – location shift | 9 | 2 | |
| GR | Increasing acute AP perturbations | 5 | 6 | 1 did not complete task, 1 stopped after 20% due to technical issues, 1 poor data quality, 3 no video data |
| | Continuous AP perturbations | 10 | 1 | |
| | Continuous ML perturbations | 9 | 2 | |
| RGR | AP perturbations (Maastricht + Hasselt) | 10 | 1 | |
| | ML perturbations (Maastricht) | 3 | 2 | |
| | AP + ML perturbations (Maastricht) | 3 | 2 | |
| | AP + ML perturbations + AST (Maastricht) | 3 | 2 | |
| | AP perturbations + AST (Hasselt) | 5 | 1 | |

PGA: Proactive Gait Adaptability; GR: Gait Robustness; RGR: Reactive Gait Recovery; AST: Auditory Stroop Test; AP: Anteroposterior; ML: Mediolateral

### **Supplemental Results**: Answers to open questions per task

All responses are translated from Dutch to English.

#### BASELINE

**Can you tell how you feel with regards to the study?**

- Open minded, genuinely curious
- ?
- I am happy to be part of it.
- Relaxed
- A little stress!
- Curious but motivated
- Tranquil
- No problem, research is necessary
- No problem
- Wait-and-see
- Fine, is important for the elderly

#### BOAT

**How did this task feel for you?**

- Well, it takes some getting used to. If I had had a short practice session, that would have been nicer
- Well, no problem. It takes a bit of getting used to
- Pleasant, was better than walking
- Pleasant
- 'Great, in the beginning no tactics that worked
- Enjoyable
- Exciting
- A lot of getting used to, especially moving the platform. It was difficult to anticipate the speed
- No problem
- Good
- Once you know how to do it, it gets easier

**What would you expect if you were to do this task as a balance training? Do you think you would get better at the task?**

- Yes, I think it will get better
- Yes
- Yes
- Yes, I suppose so
- Yes
- I think so. With repetition if you do it more.
- You'll get there in the end. Think so
- Think so
- Maybe a little

- I think it helps
- I think so

**Can you describe what you had to do?**

- Mobility, shoulder girdle, thorax
- Slalom
- Arrange steps, go faster and slower
- Steering with feet
- Better after the 2nd time. Zigzagging from left to right
- Parking a car/driving
- Around the markers, to be sent with the body, and to reach the finish line
- Steering the boat with my body
- Manoeuvring between buoys
- Steering the boat with my body
- Slalom around the arrows, 1x I went around the island, that was not good. You just have to know

**What did you think of the task?**

- Relevant
- fun
- fun
- good
- fun
- fun. Good balance is needed
- Interesting
- Very nice
- Reasonable
- fun
- Very nice, not strenuous, easy

**How did you feel during the task?**

- Focused, tension of wanting to win
- Easy
- Well, don't be afraid
- No stress,... Habituation
- Good. 2nd time was better
- Relaxed. A challenge
- Quiet
- Not good at first. Felt like I was failing, after that it got better
- Normal
- Excellent
- Good. I liked it, I felt relaxed

**Can you tell us more about how you felt with regards to nervousness, anxiety, and stress?**

- Nothing, healthy kick
- No stress, quiet environment

- Startthought she was going to fall
- Not experienced
- Not. Thought I hit the obstacle, and I'm not allowed.
- Slightly nervous
- No stress or anxiety, but an occasional adrenaline rush through the game
- Not anxious
- Nothing
- No not experienced

#### CITY RIDE

**How did this task feel for you?**

- A bit of joyriding, a bit to kick on
- Slightly more intense and interesting than the boat task
- Harder. I still had to discover how the car drove. Sometimes it went a bit fast, to the back for a little more control
- Pleasant
- Also good, seems more dangerous
- Nervous
- Bit difficult
- Intense in the sense of you want to do well but you can't
- fun
- Well strenuous
- A bit tricky, I hit the pavement and some cars

**What would you expect if you were to do this task as a balance training? Do you think you would get better at the task?**

- Yes
- Not much I think
- Little to do with balance
- Suppose so
- I think so, hard to estimate
- No, I don't think so
- I think this leads to improvement
- Think so
- Maybe
- I think so
- Yes!

**Can you describe what you had to do?**

- Reflexive, psychomotor concentration versus visual
- Dodging cars
- Guiding the car through the city, not colliding. I collided 1 time.
- Driving a car with feet by finding the right balance
- Slalom between the cars
- Try not to hit the obstacle. There could be an oncoming car or embroidery (curb)

- Dodging cars
- Driving a car with my feet and avoiding cars
- Trying to stay on the road and avoid cars
- Trying to avoid cars and get as far as possible
- To move to the right when there were cars driving left or in the middle and to the left when there were cars driving right or in the middle. Stepping forward to go faster

**What did you think of the task?**

- Highly relevant
- More pleasant than the previous one, more movement
- Fun, difficult
- Very nice
- Nice though
- Fascinating but not written for me
- Very interesting
- fun
- fun
- fun
- Fun to do, but I still have to learn it

**How did you feel during the task?**

- Motivated, decides not to clash
- Nothing special, attentive
- Very concentrating
- good
- Tense a little, feeling like you weren't doing it right
- Strained
- A bit tense
- excellent
- A bit tense
- A little more tense
- Relaxed

**Can you tell us more about how you felt with regards to nervousness, anxiety, and stress?**

- almost nothing
- Nothing
- Fear does not. A bit stressed after the collision.
- a little stress to do the right thing
- I wouldn't know
- Fear not, a bit nervous
- A bit nervous
- Not at all, just excitement from the game, but not afraid of falling
- Not anxious or stressed, but a little tense
- Not at all
- No nervousness, no fear or stress

### AST

**How did this task feel for you?**

- Accustom. Sometimes ambiguous, neither high nor low
- Good, but more tense
- Harder to concentrate
- Sometimes a bit strange, staying concentrated is difficult
- Good. A bit tense, little difference between high and low (confusing)
- Nice to meet you, it was a bit difficult the concentration
- Strenuous
- Excellent
- Okay
- Pay close attention
- Concentrate, sometimes I doubted and then I had to pay attention quickly for the next one

**What would you expect if you were to do this task as a balance training? Do you think you would get better at the task?**

- Yes undoubtedly, you have to combine stimuli
- No idea
- Think so
- Hard to say, doesn't seem to have much to do with balance
- No
- Oh yes
- Doubt
- Think it would stay the same, don't think you can train this
- No
- Think so
- Yes!

**Can you describe what you had to do?**

- Pitch recording and communication
- Walking and distinguishing between high and low sounds
- Walking and concentrating on sounds
- Tone Say (High-Low)
- High or low answers while walking. Walking + concentrating
- As I walked, the words I heard when the pitch was low say low, and high then say high.
- Walking and saying pitch out loud
- Walk straight and in the meantime pay attention to high and low tones and say them and not the word that is said
- 2 treble and bass played. Say whether it was high or low.
- Pay close attention to the tone and not to the speech
- Concentrate well on the high and low tones, if you doubt for a moment the next one will come, so you have to pay close attention. It is a good exercise for the elderly

**What did you think of the task?**

- Awakening, alerting
- Good, interesting
- Instructive to concentrate more
- Interessanr, landscape distracts
- fun
- You had to concentrate
- Interesting
- Nice though
- Okay
- difficult
- Interesting, you have to react quickly

**How did you feel during the task?**

- alert
- A little more tense
- Sometimes clumsy when she realized she was wrong
- good, slightly tense
- Relaxed
- I found it fascinating
- Quiet
- Good
- Okay
- Good, staying well concentrated, but for a musician this is fun and easier
- I felt good, you have to keep the concentration, otherwise it will go wrong with the next tone

**Can you tell us more about how you felt with regards to nervousness, anxiety, and stress?**

- Not tense. Still tension, no stress tension
- A bit tense to give the right answer
- Not nervous. Fear sometimes of answering the wrong way. No stress
- A bit nervous
- I hadn't
- Maybe a little nervous because you don't want to make a mistake
- Not at all
- Wasn't there
- Not
- Nothing
- No nervousness, no fear or stress

#### PLACEBO GENERAL

**Do you believe that participating in this training (combination of all previous tasks) could lead to an improvement in your balance?**

- Undoubtedly! But in daily life I have to study the skills.
- I honestly don't know. Studies are important. But I find it difficult to say whether it works. I think so

- I think so. If she would do this more, this would improve. Training against falls very important
- I think so
- Yes, difficult to estimate. She knows she is safe here so difficult to compare with ADL
- I think so
- Naturally
- Maybe
- I don't know
- I think so, yes
- I think it's very good for balance and reaction. All tasks have to do with reaction and concentrating

**Would you recommend this balance training to others? Why?**

- Yes, because by creating automatic you have fewer accidents and 'stupid cases'.
- Yes, if it can help, that's important
- Yes, because some have little balance and therefore drop out of walking in her walking club.
- Yes, if they need it
- Yes
- Yes, for yourself, it is a study every participation is positive and benefits
- Yes, above 65+. Especially the boat and car game, because you actually have to lean there
- I would recommend it, because you are working in a different way than usual so that you get more balance
- No idea
- I think so. It feels like you have to apply a technique to stay stable
- Yes. Good test for the elderly that I hang out with, but of course they have to be able to walk, otherwise it won't work

### PGA ASSESSMENT

**Can you describe what you had to do?**

- Stepping over white falcons, don't dodge
- Avoiding surfaces, adjusting steps
- Avoiding white areas. It was exciting whether they were going to come left or right and fast or slow
- Avoiding white blocks, not stepping on them
- Stepping over the planes
- While walking on the treadmillavoiding white obstacles and stepping over them. Small/narrow areas came more suddenly, were more difficult
- Walking and avoiding obstacles
- Walking and avoiding the white blocks
- Avoiding white areas and stepping over them
- Dodging the white areas
- Avoiding obstacles

**What did you think of the task?**

- Monotonous, attention-slackening
- No problem
- fascinating
- Very interesting
- fun
- Fascinating and strenuous
- difficult
- Was nice though
- Funny
- Quite difficult
- Very nice

**How did you feel during the task?**

- Little interested, bored
- Ordinary
- relaxed
- Good, a bit searching for the right technique
- A little tense. She didn't know if she was doing it right
- Tense, you don't want to make mistakes
- A bit nervous
- Good
- A bit tense
- A little more strenuous
- Good

**Can you tell us more about how you felt with regards to nervousness, anxiety, and stress?**

- Attention decreasing, drowsy
- No
- absent, more at ease. Only tension when the planes came quickly, difficult to adjust
- A bit nervous because it was looking for the right technique
- Not really
- A little stressed
- A little nervous, no fear
- Not at all
- Not at all
- Slightly nervous
- No stress, anxiety or nerves

### PGA STEPPING CHALLENGE

**Can you describe what you had to do?**

- On the white surfaces step and walk the colored
- Step on the white areas and avoid the red ones
- Stepping on the treadmill on the white surfaces and dodging the red ones
- Step on the white surfaces, not on red
- White areas touch, red ones don't

- While I walked with my feet on the white surfaces but not on the red ones
- Step on the white squares, and not on the red ones
- step on the white surfaces
- Step on the surfaces, not on red
- The white areas touch, red does not
- step on the whites and dodge the reds

**What did you think of the task?**

- Exciting in the good sense (task tension)
- Good
- A lot of concentration is needed for the task. There is a certain level of difficulty involved. You look at the rolling treadmill and moving surfaces, more forward is faster is harder
- Requires balance, pay close attention
- Nice, good
- fun
- Interesting
- fun
- Not fun, because I had to look down all the time
- Intense, went fast
- Very nice

**How did you feel during the task?**

- Alert
- Slightly nervous, because I had a bad start
- Concentrated, sealed on the belt
- Good, slightly tense
- Just good, relaxed
- Very relaxed
- Quiet
- good
- Not nice
- Good
- Good

**Can you tell us more about how you felt with regards to nervousness, anxiety, and stress?**

- Virtually nothing
- Not really
- Not nervous/anxious. Stress when too far forward, too fast
- A bit nervous
- I can't say anything about it. A bit nervous because you want to do it right
- No, actually quiet huh
- Not at all
- Nothing
- Tense, no fear or stress
- Nothing
- No stress, anxiety or nerves

### PGA STEPPING CHALLENGE MOVING BLOCKS

**Can you describe what you had to do?**

- Step on the white surfaces
- Walking on the white surfaces that sometimes appeared irregularly
- Step on the white surfaces
- Hitting the white areas that were more spread out
- While I was running on the treadmill I had to stand on the white obstacles
- Step on the white surfaces
- Stepping on the light surfaces
- Trying to hit the white areas
- Step on the white squares, and sometimes they were in the middle and sometimes in the same place

**What did you think of the task?**

- Well, nothing special
- Task while maintaining concentration and finding a lot of balance. Real task for balance
- A lot harder and harder to keep balance. Had to make more adjustments
- fun
- Nice task, relaxing
- Fine
- Not fun, again because you constantly have to look down
- Good to do
- Easy and fun. These are all fun tests and require concentration and movement

**How did you feel during the task?**

- Good
- Relaxing, provided you maintain concentration
- Good
- Relaxed
- Very relaxed
- Excellent
- Strained
- Enjoyable
- good

**Can you tell us more about how you felt with regards to nervousness, anxiety, and stress?**

- No stress, but attentive
- No fear, nervousness
- A bit nervous
- Not
- No, relax
- Wasn't there
- Not at all
- Not at all
- None of them

### GR ASSESSMENT

**Can you describe what you had to do?**

- Earthquake Handling
- Keeping balance as the speed of the treadmill changes
- Walking on the treadmill and sharing my experience of becoming aware of the movements of the belt
- Accelerating and slowing down the treadmill, saying that it happened and whether I had to adjust my stride. And say how stable I felt
- Treadmill running, see if you can continue walking properly after obstacle
- Maintaining balance while running on the treadmill. (Walking bent over is easier)
- Walking, with small disturbances, felt like the speed was too slow
- Describe how to adjust stable + steps
- Trying to keep walking normally
- Walking on the belt and continuing to walk even when it stopped for a moment

**What did you think of the task?**

- Relevant but monotonous
- Good
- special experience. Really an exercise against falling/feeling what happens when you fall
- Interesting
- A little less fun
- difficult and strenuous
- Good
- Okay
- Pretty difficult
- Nice, nice

**How did you feel during the task?**

- Blunted
- Good
- Concentrated walking
- Good
- Good
- Tense, you don't want to make mistakes
- Excellent
- A bit tense
- Good
- Very good

**Can you tell us more about how you felt with regards to nervousness, anxiety, and stress?**

- No stress
- No
- nothing, relax
- Slightly nervous with balance
- Not really

- A bit afraid of losing balance and you don't want to fall
- Not present
- Not at all
- Nothing
- Not bothered by that

#### GR CONTINUOUS AP

**Can you describe what you had to do?**

- Maintaining balance after treadmill wobble
- Maintaining balance at uneven speeds
- Walking on the treadmill and experiencing what the belt does and how I react to it
- Trying to maintain balance
- Walking on a treadmill with all kinds of obstacles
- Maintaining balance on the treadmill despite constant obstacles
- The treadmill made bumps that made it look like the surface was unstable
- Staying upright
- Trying to keep walking steady
- On the treadmill, which went left, right, backwards, sometimes stopped. Finding balance

**What did you think of the task?**

- Challenging, earthquake feeling
- A little more exciting, more challenging
- striking. That's how someone who has been drinking feels, I think. It sometimes held her back. Focusing on balance
- Interesting
- If I have to go for a walk like that, I'll never go again. A little less fun
- I was constantly challenged, more like challenged/bullying behavior
- Excellent
- Not fun, because of the intensity of the challenge
- Was strenuous
- Slightly more difficult than the other tests. I found this to be the most balance test of the whole

**How did you feel during the task?**

- Be careful, wary. As if the ground was sinking under their feet
- More tense, because you don't want to lose your balance
- concentrate but still
- Good
- Tense, even though she knows she can't fall
- Tense, in the positive sense
- Felt a bit like I was drunk
- Not good
- Good
- Good

**Can you tell us more about how you felt with regards to nervousness, anxiety, and stress?**

- Not, but vigilant
- A little stressed
- not nervous. No fear. Sometimes I have to get back on track
- No stress
- A little fear, a little stress
- A bit of stress, you don't want to give up. constant
- Nothing
- Not anxious
- Nothing
- No

#### GR CONTINUOUS ML

**Can you describe what you had to do?**

- Keeping balance with left-right movements
- Walking on a treadmill and constantly finding my balance
- Trying to maintain balance
- Treadmill walking with zig-zag motion
- Keeping balance during walking exercise, felt like walking on a boat
- Walking on the treadmill and it felt like the platform was shifting
- Keeping balance
- Trying to keep running as well as possible
- As if I was walking on a sloping road

**What did you think of the task?**

- Harder to keep balance
- very intensive
- Bit weird
- fun
- Actually quite nice. Someone who feels dizzy experiences it this way
- fun
- It wasn't too bad, not that extreme
- was more intensive
- Easy to do, easy

**How did you feel during the task?**

- Good
- first time there was a little bit of fear
- Good
- At ease, quiet
- Good. I thought I'm on a boat, without seasickness
- Also good, it felt like I was walking on a cloud
- Regular correction
- Good

- Good

**Can you tell us more about how you felt with regards to nervousness, anxiety, and stress?**

- No stress, but always expectation that more would happen
- not nervous or stressed. She became insecure because she didn't know what was happening. A bit anxious
- No problem. Slightly nervous maybe
- I can't answer anything. Not really present.
- Pretty relaxed
- Nothing
- Not at all
- All three do not
- No

#### RGR AP ACC

**Can you describe what you had to do?**

- Be alert not to fall
- Keeping balance while the band sometimes stopped
- Walking and the treadmill made me stumble
- Keeping balance, adapting to speed
- Walking on a treadmill with all kinds of malfunctions
- Maintaining balance on the treadmill while there were unexpected inhibitions
- Walking on the treadmill, with multiple balance challenges
- Staying on your feet
- Trying to walk as stable as possible
- The band was always braking and so I had to pay attention and lift my feet better

**What did you think of the task?**

- Sharp, relevant, lifelike
- Good
- fun
- Interesting
- A little less fun. Fear of falling
- quite strenuous
- Not so nice, I noticed that I was shocked every time
- Not nice, because you are in danger of falling
- Intense
- More intensive, I had to pay more attention than with the rest

**How did you feel during the task?**

- awake, on my guard
- Interested, attentive
- quiet
- Good
- Nervous, you don't know what's coming your way

- strained
- Very alert
- Strained
- Tense
- good

**Can you tell us more about how you felt with regards to nervousness, anxiety, and stress?**

- not stressed
- no
- nothing, not all three. Starts to get used to it. Curious when it came
- A bit nervous
- A little bit of everything
- A bit nervous
- Occasionally felt fear of falling
- A bit anxious
- Not at all
- no

#### RGR AP AST

**Can you describe what you had to do?**

- Maintaining balance and indicating tones
- Walking on the conveyor belt. I was stopped and tripped and in the meantime listening to high and low
- Maintaining balance. Say if I heard a high or low tone
- Walking on a treadmill with disturbances, distinguishing high and low tones
- Maintaining balance on the treadmill despite the obstacles while correctly indicating the high and low tone

**What did you think of the task?**

- Slightly more difficult
- Paying attention to two things very intensively and concentrated. Correct steps and tones
- Interesting
- A little less fun, because you have to concentrate more on 2 things
- Fascinating and strenuous

**How did you feel during the task?**

- A little more tense
- A bit anxious. "Oops"
- Good
- A little nervous
- concentrated

**Can you tell us more about how you felt with regards to nervousness, anxiety, and stress?**

- No stress

- Not nervous, but feeling stress and anxiety coming up
- Slightly nervous
- A bit nervous, no fear
- rather tense not to make mistakes

#### RGR ML

**Can you describe what you had to do?**

- Walking on the treadmill,and the platform moved
- Staying upright
- Trying to keep walking as straight as possible

**What did you think of the task?**

- In the beginning I was shocked, but not later
- Not nice, same reason as last one
- difficult

**How did you feel during the task?**

- Excellent
- Strained
- Excellent

**Can you tell us more about how you felt with regards to nervousness, anxiety, and stress?**

- It was a bit of a shock in the beginning, less so later
- A bit anxious
- Nothing

#### RGR AP + ML

**Can you describe what you had to do?**

- Walking on the treadmill, and there were several ways in which the treadmill changed
- Balance of the elderly
- Trying to walk steadily

**What did you think of the task?**

- Unpredictable
- Less bad than the previous one (correct see comments)
- Was doable

**How did you feel during the task?**

- Alert
- Pretty okay
- Good

**Can you tell us more about how you felt with regards to nervousness, anxiety, and stress?**

- Little fear

- A bit anxious
- Nothing

#### RGR AP + ML + STROOP

**Can you describe what you had to do?**

- Do two things, walk and listen to the pitch. And making sure I stayed stable. So actually three things. More complex I had to pay close attention That didn't play a role
- Keeping balance + indicating pitch. Not really difficult but occasionally losing attention Reasonably okay Not at all
- Walking stably and indicating the pitch. Nice Fine Not at all

**What did you think of the task?**

- More complex
- Not really difficult, but occasionally lost attention
- Fun

**How did you feel during the task?**

- I had to pay close attention
- Pretty okay
- Fine

**Can you tell us more about how you felt with regards to nervousness, anxiety, and stress?**

- That did not play a role
- Not at all
- Not at all
